# Background covariance adjustment distills shared genetic architecture across neurodevelopmental and neurodegenerative disorders

**DOI:** 10.64898/2026.03.08.26347891

**Authors:** Xinwen Huang, Yunpeng Wang, Qingyuan Zhao, Zijun Gao

## Abstract

GWAS increasingly reveal shared genetic influences across neurodevelopmental, psychiatric, and neurodegenerative traits. However, cross-trait genetic covariance derived from GWAS summary statistics can be inflated by sample overlap and other structured background effects, obscuring higher-order genetic organization. We extend PathGPS, a recently developed statistical method that estimates an adjusted genetic covariance by subtracting a background covariance learned from weakly associated variants, and then extracts reproducible low-rank structure using rotation and bootstrap aggregation. When applying to 15 phenotypes related to neurodevelopmental and neurodegenerative disorders, the adjusted analysis yields four stable clusters with an interpretable topology. Adjusting for background covariance, which appears to be related to traumatic life experiences, sharpens the cluster boundaries and substantially shifts the clustering result for post-traumatic syndrome disorder. Simulations with controlled overlap and structured background covariance show that PathGPS has improved factor recovery relative to substantially shifts the clustering result for post-traumatic syndrome disorder.

## Introduction

Large-scale genome-wide association studies (GWAS) of neurodevelopmental, psychiatric, and neurodegenerative phenotypes increasingly indicate that genetic risk is not confined within traditional nosological boundaries but broadly shared across different diagnoses^1-5^. Recent cross-disorder analyses show pervasive pleiotropy, with many loci contributing to risk across multiple conditions, and with multivariate structure that only partially aligns with clinical categories. As GWAS sample sizes continue to grow, the key challenge has shifted from establishing heritability within individual disorders to organizing shared genetic architecture across many traits in a way that is statistically principled, replicable, and biologically interpretable.

At the same time, cross-trait similarity among brain-related phenotypes reflects more than shared genetic liability. Epidemiological evidence documents substantial clustering of exposures and contextual risk factors—such as trauma burden, socioeconomic adversity, and correlated health behaviours—that contribute to comorbidity and may induce correlated outcomes across disorders and traits^6-8^. In genetic studies, additional sources of shared background structure can arise from overlapping cohorts, correlated ascertainment, participation and selection mechanisms, and measurement or diagnostic artifacts. Recent work further emphasizes that large-scale “shallow” phenotyping (for example, self-report questionnaires or EHR-based codes) can incorporate heritable confounders—such as response bias and selective participation—into case definitions, generating shared covariance in summary statistics that can masquerade as shared etiology across disorders^9^. When multi-trait analyses are built from heterogeneous meta-analytic GWAS and partially overlapping samples, these shared background effects can propagate into cross-trait covariance in marginal association statistics, potentially blurring higher-order genetic organization even when individual-trait summary statistics are well-powered. Distinguishing robust pleiotropic genetic structure from background cross-trait covariance is therefore central to building interpretable maps of shared genetic architecture.

Most widely used methods for multi-trait analyses begin with pairwise relationships—estimating genetic correlations (often using linkage disequilibrium score regression, LDSC) and then applying downstream models such as Genomic structural equation modelling (GenomicSEM) or exploratory factor analysis on the resulting trait–trait genetic correlation matrix^10,11^. Such pipelines are powerful and computationally convenient, but they compress the full SNP-by-trait summary-statistics matrix into a set of bivariate summaries, which can limit sensitivity to higher-order relationships that are only apparent when traits are modeled jointly. Moreover, while methods such as LDSC and GenomicSEM can incorporate sampling covariance and account for overlap-related effects under their modeling assumptions, in practice multi-trait panels may exhibit structured background covariance that is difficult to diagnose and that can attenuate factor separation, reduce interpretability, and inflate apparent connectivity between otherwise distinct domains. Complementary multivariate approaches, such as Genetic Factor Analysis (GFA), operate more directly on the standardized GWAS effect matrix and explicitly estimate background correlation structure, including components that may reflect overlap or other shared background effects^12^. However, a practical gap remains: in real multi-trait neuropsychiatric settings, background cross-trait covariance can be structured and trait-specific, and its impact on inferred modules is rarely explicitly characterized, stress-tested, and compared against a corrected alternative within a unified workflow.

Here we build on PathGPS, a multivariate framework that infers shared genetic architecture from SNP-by-trait co-appearance patterns in GWAS summary data^13^. PathGPS models shared genetic architecture using latent mediators, and uses repeated subsampling and rotation to recover stable, interpretable multi-trait components that go beyond pairwise correlation structure. In this work, we extend PathGPS with an explicit background covariance adjustment designed for heterogeneous multi-trait GWAS panels. The key idea is to estimate a shared background covariance component from variants showing little evidence of association across the analyzed traits (weak-association variants), and to subtract a scaled version of this background covariance from the covariance computed from associated variants to obtain an adjusted cross-trait covariance that better reflects genetically mediated structure. We refer to this estimated background component as a shared residual (or shared background) covariance (SRC) term that captures any structured cross-trait covariance in the summary statistics that is not attributable to the targeted genetic signal, including contributions from sample overlap, ascertainment and selection mechanisms, correlated non-genetic exposures, measurement artifacts, and heritable confounding arising from shallow phenotyping and selective participation that can induce shared, off-target signal across disorders, and potentially diffuse polygenic background under thresholding.

We couple this adjustment with a stability-focused implementation—bootstrap aggregation, trait–trait co-appearance matrices, and consensus clustering—and with permutation-based tests that quantify whether the resulting modular structure exceeds that expected under label-randomized null structure. We then apply the framework to a harmonized compendium of 15 brain-related phenotypes spanning neurodevelopmental conditions, major psychiatric disorders, cognition-related traits, and neurodegenerative diseases, and benchmark performance in simulations designed to mirror realistic overlap and structured background covariance.Together, this work provides a practical and reproducible approach to distilling shared genetic architecture in the presence of structured cross-trait background effects, and to producing multivariate trait modules that can be compared directly with and without background covariance adjustment.

## Results

### PathGPS overview and the SRC-adjusted covariance

We extended PathGPS to operate directly on a harmonized SNP-by-trait summary-statistics matrix and to explicitly remove structured cross-trait background covariance prior to module discovery (Figure 1). In brief, PathGPS treats the standardized genome-phenome association matrix as the observable output of (i) shared genetic mediators that induce structured pleiotropy and (ii) additional cross-trait covariance that is not captured by the targeted genetic signal (*e*.*g*., arising from sample overlap, ascertainment, participation/selection, measurement artifacts, or other shared background effects). Operationally, we estimate this second background component using variants with little evidence of association across the trait panel and refer to it as a shared background covariance (SRC) component.

**Figure 1.**
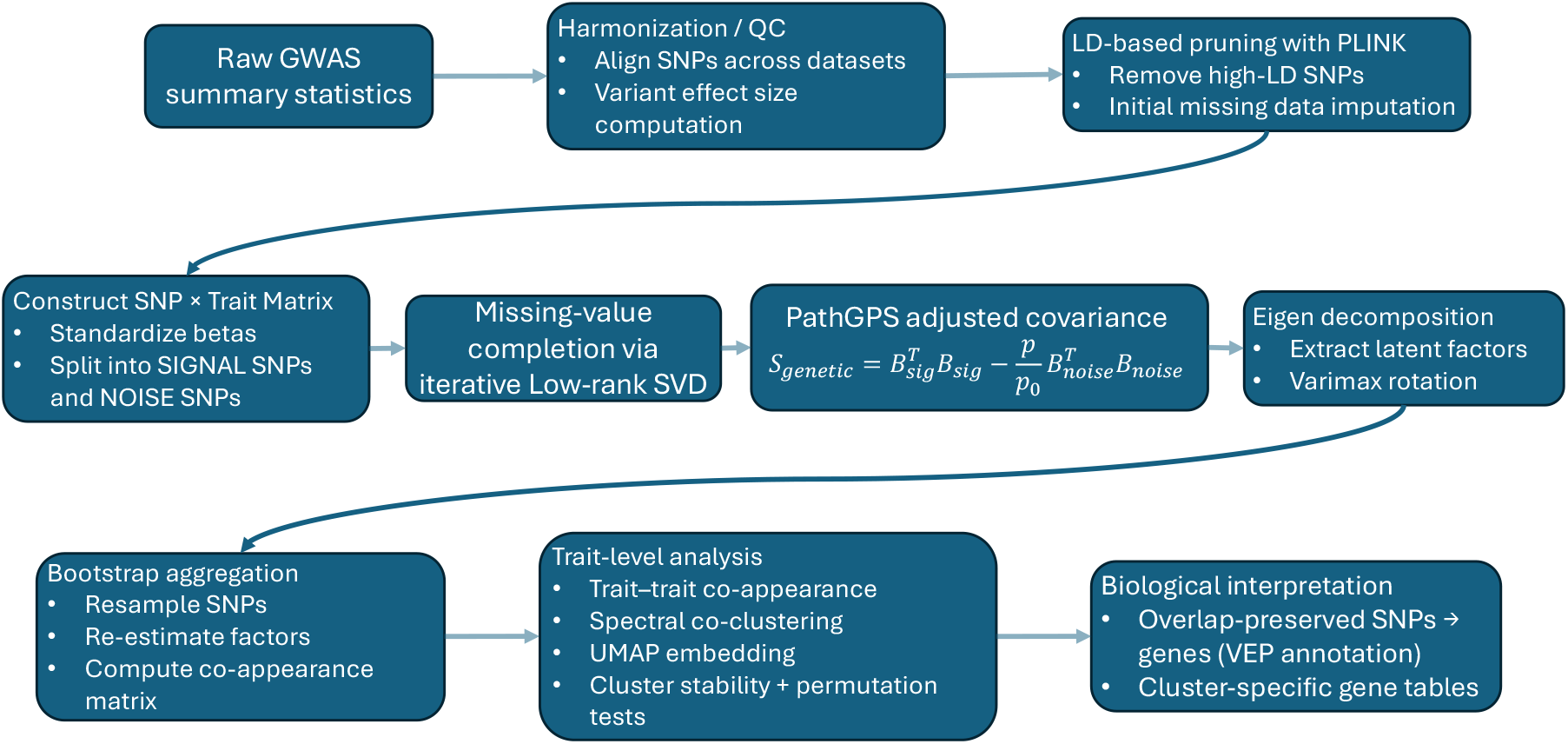
Follow-chart for PathGPS analysis. Abbreviation used: GWAS, genome-wide association studies; QC, quality control; SNP, single nucleotide polymorphism; LD, linkage-disequilibrium; SVD, singular value decomposition.

Given a set of “signal” variants S and “weak-association/noise” variants N (defined below for each analysis), we compute trait–trait covariance matrices from the corresponding standardized marginal associations:

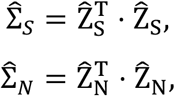

and define an SRC-adjusted covariance estimate as

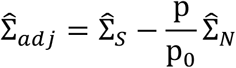

Where *p* is the number of signal SNPs, and *p*_0_ is the number of noise SNPs.

We then extract low-rank structure from 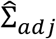 via eigen-decomposition (retaining the top r components), apply varimax rotation to improve interpretability, and use repeated SNP subsampling (bootstrap aggregation) to quantify stability of the inferred structure.

To summarize reproducible multi-trait organization, we aggregate results across bootstrap runs into a trait–trait co-appearance matrix C, where C_*ij*_ records how frequently traits *i* and j are assigned to the same latent component across runs. We then apply spectral clustering^14^ to C to define consensus trait modules and visualize the resulting trait geometry using Uniform Manifold Approximation and Projection (UMAP)^15^ applied to the consensus similarity structure.

Finally, we evaluate whether the inferred modular organization exceeds that expected under a label-randomized null. Specifically, we compute (i) a global contrast statistic comparing within-module vs across-module co-appearance and (ii) cluster-specific contrast statistics comparing within-module co-appearance to co-appearance between that module and the remainder of traits (**Methods**). Empirical cluster discrepancy scores are obtained by permuting trait labels while preserving module sizes.

### PathGPS improves recovery of latent genetic factors in simulations with overlap and structured background covariance

We first benchmarked PathGPS in simulations designed to mirror two major challenges in real multi-trait GWAS: sample overlap and structured shared background variance (i.e., correlated non-genetic components) (details see **Methods**). In the data-generating model, traits arise from a low-dimensional genetic mediator structure plus an additional correlated background component that induces cross-trait covariance not attributable to the genetic mediators. We varied the strength and rank of this background component, spanning settings from negligible overlap/no structured background covariance to strongly overlapping designs with pronounced low-rank residual structure.

We compared SRC-adjusted PathGPS against six baselines including several matrix decomposition and correlation-matrix factorization approaches: 1) singular value decomposition (SVD), singular value decomposition and Hilbert transform (SVD-HT), 2) sparse principal components (SPC), 3) maximum-likelihood factor analysis applied to an LDSC-like genetic correlation matrix with four and eight factors (MLFA_4 and MLFA_8; representing a GenomicSEM-style exploratory factor analysis baseline)^11^, and 4) Genetic Factor Analysis (GFA) ^12^. We evaluated each method by its ability to recover the ground-truth latent factor structure and avoid extraneous (spurious) factors (**Figure 2**; **Methods**).

**Figure 2.**
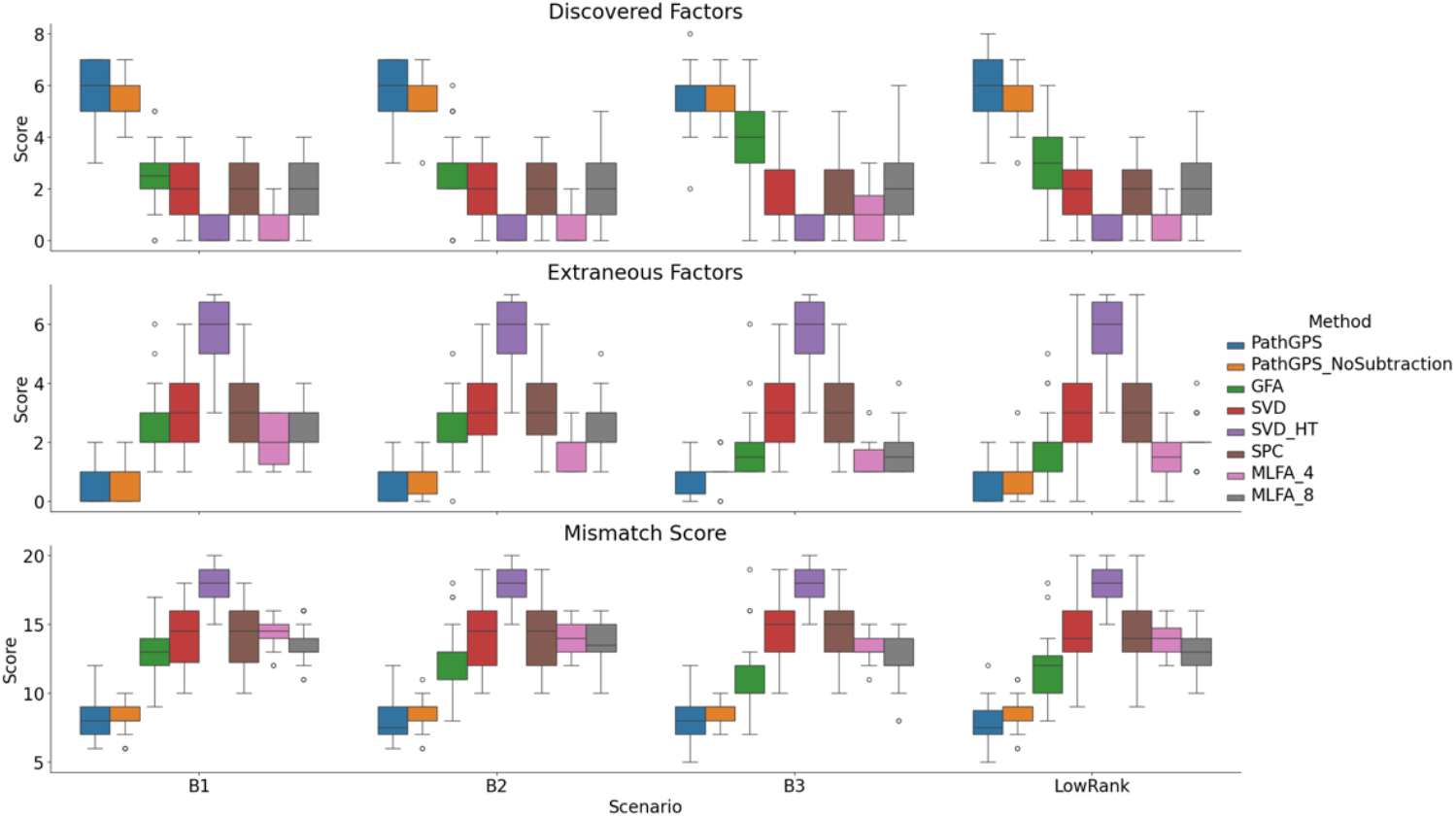
Performance of PathGPS compared with baseline methods across simulation scenarios. Boxplots summarize distributions of discovered factors, extraneous factors and mismatch scores across 30 replicates for each scenario (B1-B3, and LowRank). Only results for the 30-trait, eight-factor model settings are shown here; other simulation scenarios are presented in **Supplementary Note** and **Supplementary Figure 2** and **3**.

Across simulation settings, SRC-adjusted PathGPS achieved the lowest mismatch to the true latent factors, with the advantage most pronounced in regimes where sample overlap and structured background covariance were strong (**Figure 2**; **Supplementary Figs. 2** and **3**). PathGPS without SRC subtraction remained competitive and generally outperformed the LDSC-based factor-analysis baselines, while GFA performed better than SVD/SPC but was less accurate than PathGPS in the most confounded settings. Collectively, these results support the premise that explicitly attenuating structured background covariance—rather than learning latent factors from total covariance—can improve isolation of genetically mediated multi-trait structure when overlap and shared residual effects are present.

### SRC adjustment steepens and deconfounds the low-rank spectrum and identifies a dominant background axis

We next applied PathGPS to a harmonized set of GWAS summary statistics spanning 15 brain-related phenotypes (**Methods** and **Supplementary Table 3**), including neurodevelopmental and psychiatric disorders (*e*.*g*., attention deficit hyperactivity disorder, autism spectrum disorder, schizophrenia, bipolar disorder, post-traumatic stress disorder, major depressive disorder, panic disorder, obsessive–compulsive disorder/obsessive–compulsive symptoms, anorexia), personality/cognitive traits (neuroticism, educational attainment), and neurodegenerative outcomes (Alzheimer’s disease, Alzheimer’s disease and related dementias, amyotrophic lateral sclerosis). For this analysis, we defined signal variants as SNPs with *p* < 5 × 10^−3^ in at least one trait and weak-association/noise variants as SNPs with *p* > 5 × 10^−2^ across all traits, yielding the two SNP sets used to compute 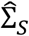 and 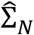.

SRC adjustment materially altered the trait covariance spectrum (Figure 3). In the unadjusted analysis, the leading components showed comparatively slower singular-value decay, consistent with broad shared covariance inflating apparent connectivity across traits. After SRC subtraction, the singular values decayed more steeply, and the adjusted spectrum exhibited an elbow after approximately the first four components (Figure 3), motivating our focus on a four-component representation for downstream module construction in this panel.

**Figure 3.**
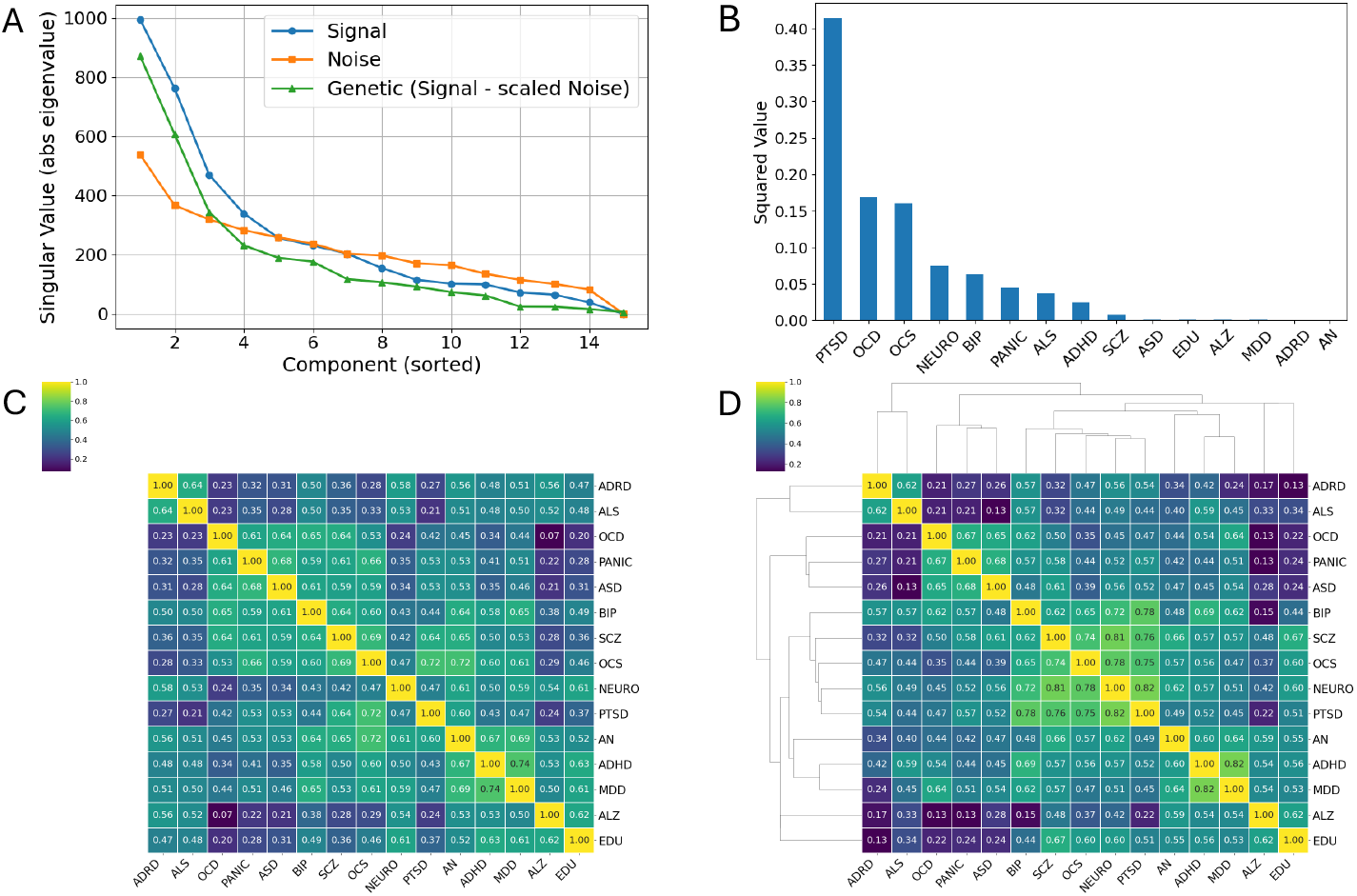
SRC adjustment steepens the trait covariance spectrum and sharpens consensus co-appearance structure. A. Singular value spectrum of the trait–trait covariance estimated from signal variants and the SRC-adjusted covariance, showing a steeper decay after background subtraction and an elbow at approximately four components in the 15-trait panel. B, Trait loadings on the leading eigenvector of the weak-association/noise covariance, highlighting a dominant shared background axis driven by PTSD (loading = 0.64). C,D, Consensus trait–trait co-appearance matrices summarizing bootstrap aggregation of PathGPS without SRC adjustment (C) and with SRC adjustment (d). Each entry denotes the fraction of bootstrap runs in which traits i and j are assigned to the same latent component (values range from 0 to 1). Abbreviations: SRC, shared residual/background covariance; PTSD, post-traumatic stress disorder.

To characterize which traits contributed most strongly to the weak-association covariance, we examined trait loadings on the leading eigenvectors of 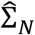. The top eigenvector of the noise/weak-association covariance captured a broad SRC axis (Figure 3) and was dominated by PTSD (loading 0.64), indicating that—within this panel—PTSD contributes disproportionately to structured background covariance as captured by weakly associated variants. This observation is consistent with the possibility that PTSD’s cross-trait similarity in summary statistics is particularly sensitive to cohort composition, ascertainment, and exposure-linked structure (while not implying that SRC is purely environmental)^16-18^.

### SRC subtraction sharpens consensus co-appearance structure and increases modular coherence

We then evaluated whether SRC adjustment improves the clarity and reproducibility of trait modules inferred by PathGPS. Without SRC subtraction, the trait–trait co-appearance matrix showed diffuse structure: emergent groupings were present but boundaries were blurred, and a subset of traits exhibited elevated co-appearance across multiple putative modules (**Figure 3**). After SRC adjustment, co-appearance structure became visibly sharper, with higher within-module similarity and reduced cross-module co-appearance (**Figure 3**).

Permutation testing corroborated this improvement. SRC adjustment increased the global contrast statistic from *T*_obs_ = **0.1681**(unadjusted) to *T*_obs_ = 0.2059(SRC-adjusted), and the global modular organization was significant under permutation (p = 0.001). In addition, a larger fraction of module-specific contrasts reached conventional significance thresholds (permutation p < 0.05), indicating that the improved module boundaries were not driven by isolated trait pairs but reflected a broader increase in within-module coherence.

Notably, SRC adjustment also altered the placement of PTSD in the consensus clustering. In the unadjusted analysis, PTSD behaved as a weakly connected singleton (**Figure 3**). After SRC subtraction, PTSD clustered with schizophrenia and bipolar disorder (**Figure 3**), suggesting that attenuating shared background covariance can reveal a module-specific genetic covariance component aligning PTSD with the psychosis–bipolar axis within this multivariate representation^19,20,21^.

### PathGPS identifies four stable modules spanning neurodevelopmental to neurodegenerative phenotypes

Using spectral clustering on the consensus co-appearance matrix, SRC-adjusted PathGPS recovered four stable modules across the 15 traits (**Figure 4**). At a coarse level, these modules separated: (i) a broad neurodevelopmental/psychotic spectrum (including ADHD, educational attainment, SCZ and BIP, with bridge phenotypes), (ii) an affective–anxiety–compulsive module (including MDD, panic disorder, ASD and OCD-related phenotypes), and (iii–iv) two neurodegeneration-related groupings that separated Alzheimer’s disease and related dementias/ALS from Alzheimer’s disease as represented in the analyzed GWAS. A UMAP embedding constructed from the consensus similarity structure arranged traits along a continuous geometry spanning neurodevelopmental and adult psychiatric phenotypes through to neurodegenerative outcomes, with intermediate “bridge” traits occupying positions between these extremes (Figure 4). The resulting geometry and module-level structure were consistent with the presence of widespread pleiotropy together with partially separable domains of shared risk.

**Figure 4.**
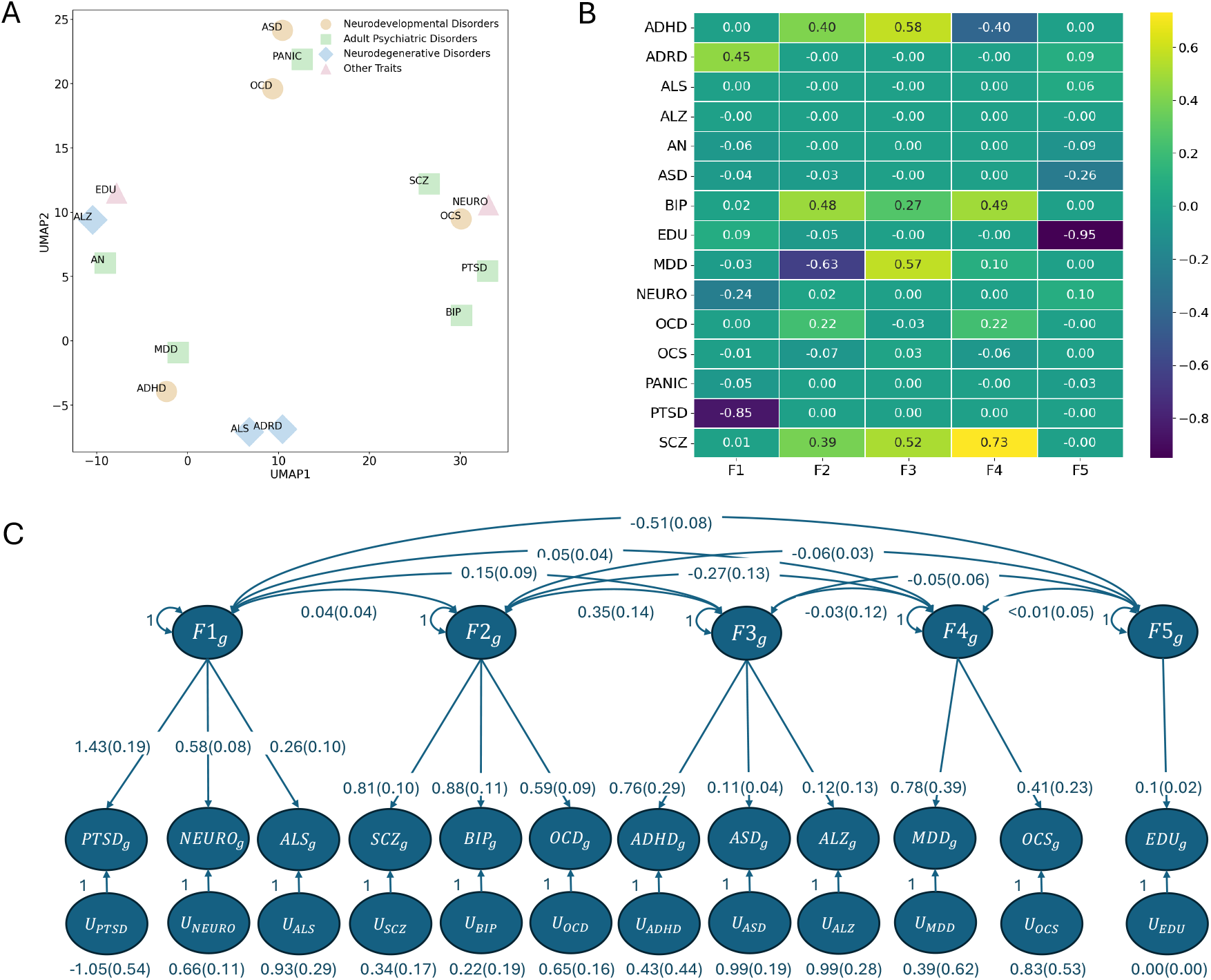
PathGPS trait geometry and comparison to GFA and GenomicSEM maps. A, UMAP embedding constructed from the normalized trait–trait co-appearance (consensus similarity) structure obtained by bootstrap-aggregated PathGPS. Each point is a trait; proximity reflects how frequently traits co-appear in the same latent components across bootstrap runs. Marker type denotes broad phenotype class (neurodevelopmental disorders, adult psychiatric disorders, neurodegenerative disorders, and other traits, as indicated). B, Trait–factor loading matrix from Genetic Factor Analysis (GFA) fit to the SNP×trait z-score matrix for the same trait panel (five-factor solution, F1–F5). Cell values show estimated trait loadings on each factor; color encodes loading magnitude and sign. C, GenomicSEM five-factor confirmatory genome-wide factor model for the retained traits. Latent genetic factors (F1g–F5g) are allowed to correlate (curved double-headed arrows; estimates with standard errors in parentheses). Directed edges denote standardized factor loadings onto trait-specific genetic components (suffix “_g”; estimates with standard errors in parentheses). U nodes denote trait-specific residual (uniqueness) components, with estimated residual variances shown beneath each U node; latent factor variances were fixed to 1 for identification.

We also interpreted these results in light of known study-design features. In particular, the AD summary statistics used here were derived from a UK Biobank family-history (GWAS-by-proxy) phenotype, which includes large numbers of proxy cases and is known to be susceptible to participation and survival-related biases^22^. Such biases can induce correlations with education-linked traits and may help explain why the AD-by-proxy phenotype does not behave identically to clinically ascertained dementia outcomes in multivariate genetic structure learning. In contrast, the clinically ascertained ADRD dataset clustered with ALS, consistent with reported cross-neurodegenerative pleiotropy, while acknowledging that the magnitude and loci of overlap vary across studies^23^.

To evaluate the stability of the inferred trait clusters, we repeated PathGPS biclustering across multiple independent runs and additionally introduced an unrelated negative-control trait (hair color; Supplementary Notes). Cluster assignments were highly consistent across runs and were not materially altered by inclusion of the negative control (Supplementary Fig. 1 and Supplementary Table 2).

### Comparison to GFA and GenomicSEM highlights differences between multivariate and bivariate maps

We next compared PathGPS modules to structures obtained from two widely used multivariate approaches: GFA (a multivariate sparse factor model) and GenomicSEM-style modeling based on pairwise genetic correlations from LDSC. While both baselines yielded a five-cluster organization in this panel (Figure 4), trait assignments differed substantially from PathGPS and from each other. For example, educational attainment loaded primarily on a single factor in the GenomicSEM analysis (model fit CFI = 0.96, SRMR = 0.14), whereas in the GFA solution educational attainment shared a factor with ASD. SCZ and BIP clustered together in both baselines, but GFA additionally placed MDD and ADHD within that factor, while GenomicSEM assigned OCD to the SCZ–BIP factor. Neither baseline placed PTSD with SCZ/BIP in this comparison, consistent with the possibility that SRC adjustment alters higher-order structure beyond what is recoverable from pairwise correlation maps.

## Discussion

In this study we extend PathGPS into a summary-statistics framework for organizing shared genetic architecture across heterogeneous brain-related phenotypes while explicitly accounting for structured cross-trait background covariance. Across simulations and a 15-trait compendium spanning neurodevelopmental, psychiatric, cognition-related, and neurodegenerative phenotypes, PathGPS recovered stable, low-rank trait organization and produced consensus modules supported by permutation testing. These results reinforce two complementary hypotheses. First, genetic sharing across brain-related phenotypes is widespread and only partially aligned with diagnostic boundaries. Second, the apparent multivariate organization inferred from GWAS summary statistics can depend materially on whether structured background covariance is attenuated prior to extracting higher-order structure.

A central practical feature of PathGPS is that it does not treat pairwise genetic correlations as the primary object for multivariate inference. Instead, it leverages the SNP-by-trait association matrix to infer latent structure from multi-trait co-appearance patterns and then evaluates reproducibility through bootstrap aggregation. This matrix-first viewpoint is useful in settings where higher-order relationships—such as bridge phenotypes that connect partially separable domains—may not be well summarized by any single pairwise metric. In the present analyses, this approach produced consensus co-appearance structure with stronger within-module coherence than pairwise-derived maps under a matched permutation-testing procedure, supporting the utility of joint multivariate inference when mapping shared architecture across many traits.

The second methodological element—explicit adjustment for shared background covariance—has the greatest interpretational importance. Operationally, we estimate a trait–trait covariance component from variants showing little evidence of association across the analyzed traits and subtract this background estimate from the covariance computed from associated variants, yielding an adjusted covariance intended to more directly reflect genetically mediated cross-trait structure. We refer to the subtracted component as a shared residual/background covariance term, emphasizing that it is not uniquely interpretable as “environmental” covariance. Rather, it can incorporate contributions from sample overlap, correlated ascertainment and selection mechanisms, participation bias, measurement artifacts, and potentially diffuse polygenic background under thresholding. The contribution of PathGPS is therefore not a claim that the estimated background term corresponds to a specific non-genetic mechanism, but that explicitly estimating and attenuating structured background covariance can clarify multivariate genetic organization in realistic multi-trait panels.

Empirically, background adjustment sharpened the modular structure recovered in the 15-trait panel and altered the placement of PTSD, which clustered with schizophrenia and bipolar disorder after adjustment in this dataset. We interpret this primarily as an SRC-sensitivity result: attenuating background covariance revealed a residual-corrected covariance pattern that aligns PTSD more closely with the psychosis–bipolar axis within this multivariate representation. This observation is consistent with prior reports of genetic overlap between PTSD and schizophrenia, while also remaining compatible with broader cross-disorder factor mappings in which PTSD often lies closer to internalizing spectra^16,18,24^. Accordingly, our results should not be read as reclassifying PTSD, but as illustrating that PTSD’s apparent cross-trait similarity is particularly sensitive to structured background covariance in heterogeneous GWAS panels, and that multivariate placement may shift depending on how such covariance is modeled.

PathGPS complements emerging multivariate methods that also operate on summary-statistics matrices. For example, Genetic Factor Analysis (GFA) models the standardized GWAS association matrix using sparse latent factors and incorporates background correlation structure that can reflect overlap-related effects. In this sense, both GFA and PathGPS acknowledge that weakly associated variants contain information about cross-trait background structure. PathGPS differs primarily in emphasis and outputs: by using a signal/background partition to form an adjusted covariance and by converting repeated decompositions into consensus co-appearance structure, it produces stability-validated trait modules and embeddings that can be compared directly with and without background adjustment. In simulations that explicitly varied overlap and structured background covariance, this adjustment improved recovery of the ground-truth latent factors relative to baselines, suggesting that subtraction-based attenuation is particularly beneficial when the background component is structured and influential. PathGPS is also complementary to other latent representation approaches^25^ that decompose summary-statistics matrices and then post-process factors for interpretability; our contribution is an explicit mechanism for quantifying and attenuating shared background covariance and for translating latent structure into reproducible modules using stability and permutation-based evidence. In addition, PathGPS introduces a bootstrap aggregation procedure that stabilizes cluster assignments across SNP resamples and summarizes uncertainty via co-appearance consensus maps.

Several limitations warrant emphasis. First, estimation of background covariance depends on how “signal” and “weak-association” variant sets are defined; while p-value thresholding provides a transparent operational definition, the optimal thresholds may vary with trait architecture, sample size heterogeneity, and panel composition. Sensitivity analyses over threshold choices—and diagnostic checks on the stability of inferred modules—are therefore important for robust application. Second, practical harmonization steps such as LD pruning, missingness handling, and low-rank completion introduce approximations that can, in principle, influence cross-trait structure; robustness analyses that restrict to SNP intersections or vary imputation/completion choices are valuable safeguards, particularly in highly heterogeneous panels. Third, the background covariance term should not be over-interpreted mechanistically without external validation (e.g., explicit sample-overlap metadata, cohort ascertainment characteristics, or independent proxies for shared exposures). Finally, our analyses are restricted to GWAS summary statistics from European-ancestry cohorts as reported by the contributing studies; extending these analyses to multi-ancestry settings will require careful harmonization and attention to ancestry-specific LD and architecture.

In summary, PathGPS provides a practical framework for organizing shared genetic architecture across brain-related traits when pleiotropy is pervasive and cross-trait background covariance is structured. By moving beyond pairwise summaries, incorporating stability via bootstrap aggregation, and explicitly attenuating shared background covariance using weak-association variants, PathGPS yields more interpretable multivariate structure and highlights phenotypes—such as PTSD— whose multivariate placement can be sensitive to background covariance modeling. This framework can serve as a foundation for downstream biological analyses that annotate module-specific loci and pathways, and for broader applications to multi-trait panels where overlap and background structure are unavoidable features of modern GWAS.

## Methods

### Study Overview

We developed PathGPS, a structural equation–based statistical framework for identifying shared genetic pathways across psychiatric, personality, cognitive, and neurodegenerative traits by directly modeling genome-wide SNP–by–trait summary-statistics matrices. Rather than focusing on single-disorder association signals, PathGPS leverages cross-trait covariance among genetic effects to uncover latent biological dimensions that organize the genetic architecture of mental health.

The generative modeling assumptions and theoretical foundations of PathGPS have been described previously^13^. In the present work, we extend the original framework in three key ways: (i) by implementing an extensive bootstrap aggregation strategy to stabilize clustering assignments, (ii) by introducing empirical permutation-based significance testing at both global and cluster-specific levels, and (iii) by incorporating robustness checks via negative controls. Figure 1 provides an overview of the full analytical pipeline as implemented in the publicly available pathgps package (https://github.com/ZijunGao/PathGPS).

All analyses were performed on harmonized GWAS summary statistics from European-ancestry cohorts.

### GWAS data sources and harmonization

We assembled publicly available GWAS summary statistics for major psychiatric disorders, personality traits, cognitive outcomes, and neurodegenerative diseases (**Supplementary Table 3**). Each dataset contained SNP-level summary statistics including chromosomal position, effective sample size, Z-statistic, p-value, and either effect sizes (BETA) or odds ratios (OR).

When standard errors were not explicitly reported, they were computed as

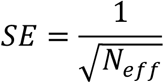

When odds ratios were provided without effect sizes, log-transformed effect sizes were computed as

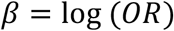

which ensures a common linear scale across binary and continuous traits.

Importantly, PathGPS does not rely on effect directionality or allele matching across studies; only the magnitude and covariance structure of SNP–trait associations are used. As a result, our procedure does not require genome build alignment or full allele harmonization, substantially reducing preprocessing requirements and reliance on study-specific metadata.

All traits were merged into a unified SNP–trait matrix using shared rs identifiers and genomic coordinates (chromosome and base-pair position), yielding a sparse matrix reflecting heterogeneous SNP coverage across studies.

All GWAS summary statistics were obtained from the original study consortia or curated public repositories (**Data availability**). For each phenotype, we used the most recent European-ancestry meta-analysis available at the time of analysis. Educational attainment results exclude participants from 23andMe, consistent with consortium data-use agreements.

### Linkage disequilibrium pruning and SNP selection

To reduce redundancy due to correlated variants, linkage disequilibrium (LD) pruning and LD estimation were performed using PLINK (v1.9.0-b.7.11, 64-bit, 19 Aug 2025)^26^. Approximately independent SNPs were identified using sliding windows of 1,000 kb with a step size of five SNPs and an LD threshold of *r*^2^ = 0.2.

Following pruning, SNPs were filtered based on cross-trait significance patterns. Variants showing moderate evidence of association (defined as p-values below the 10th percentile for at least one trait) were retained for downstream modeling. This step enriched the dataset for variants likely to contribute to shared genetic structure while preserving polygenic signal.

### LD-based imputation of missing summary statistics

Because not all SNPs were present across GWAS datasets, we implemented an LD-based donor–target imputation procedure. SNPs with missing values in fewer than eight traits were defined as light-missing targets, while SNPs with higher missingness served as potential donors.

Using PLINK-derived LD pairs, missing summary statistics were imputed by transferring trait-specific values from the most strongly correlated donor SNP (*r*^2^ > 0.4). Each donor SNP was used only once per trait to prevent propagation bias.Duplicate SNP–trait entries were collapsed by retaining the first non-missing observation. Implementation details and code are provided in the PathGPS GitHub repository.

After LD-based imputation, overall missingness across traits was substantially reduced. However, because PathGPS requires complete matrices for covariance estimation, an additional completion step was applied as described below.

### Signal–noise decomposition

To isolate shared genetic signals from background variations, SNPs were partitioned into signal and noise sets based on trait-wide association patterns. SNPs were classified as signal variants if they exhibited p-values below 5 × 10^−3^ in at least one trait, and as noise variants if all observed p-values exceeded 5 × 10^−2^. Missing p-values were skipped when computing minimum p-values for this classification.

This separation enabled estimation of trait covariance attributable to genuine genetic effects while accounting for background correlations arising from sampling variability.

### Low-rank SVD completion of signal and noise matrices

Let 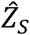 and 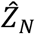 denote standardized SNP effect-size matrices for signal and noise variants, respectively. Remaining missing values in each matrix were completed separately using iterative low-rank singular value decomposition (SVD), following established matrix completion algorithms (e.g., soft-impute–style alternating minimization).

Since the rank should less than the number of traits, for each matrix, we set the rank to *r* = min(#*SNPs*, #*traits*) − 1. LD pruning and donor-based imputation substantially reduced missingness, but complete matrices are required for PathGPS covariance estimation; therefore, SVD completion was deployed as a final step.

### Structural equation model and adjusted genetic covariance

PathGPS adopts a linear structural equation model linking SNPs to traits through latent genetic and environmental mediators. Let *X* = (*X*_1_, …, *X*_*p*_) denote SNPs and

*Y* = (*Y*_1_, …, *Y*_*q*_) denote traits. We assume traits are influenced by latent genetic mediators *M* = (*M*_1_, …, *M*_*r*_) and unobserved environmental mediators *m* = (*m*_1_, …, *m*_*s*_), such that

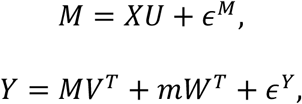

Where *U* ∈ *R*^*p*×*r*^, *V* ∈ *R*^*q*×*r*^, *W* ∈ *R*^*q*×*s*^ are coefficient matrices, and *ϵ*^*M*^ ∈ *R*^*r*^, *ϵ*^Y^ ∈ *R*^*q*^ denote zero-mean errors in the genetic mediators *M* and traits *Y*, respectively. As shown in Supplementary Section 1, the marginal SNP–trait associations satisfy

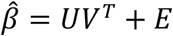

where *E* is a zero-mean residual matrix reflecting environmental mediators, sampling variation, and other non-genetic sources. We interpret this background covariance as a shared background covariance (SRC) component.

To isolate genetic structure, we estimated trait-level covariance matrices for signal and noise SNPs:

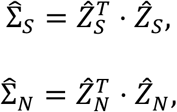

and constructed the adjusted genetic covariance

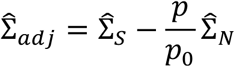

where *p* and *p*_0_represent the number of signal and noise SNPs, respectively.

The expectation of this difference matrix is positive semidefinite. In practice, small negative eigenvalues may arise from sampling variability; however, these are typically negligible compared to the leading eigenvalues of interest. Because PathGPS focuses exclusively on dominant components, we do not explicitly enforce positive semidefiniteness. For numerical stability, SVD can be used interchangeably with eigen decomposition, yielding equivalent leading components.

### Latent pathway extraction and rotation

We estimated the column space of *V* using the leading eigenvectors (or singular vectors) of 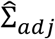. The number of retained components was selected based on the eigenvalue spectrum, which consistently showed an inflection around five components.

To enhance interpretability, we applied varimax rotation, producing sparse and interpretable trait loadings. Each latent pathway represents a coordinated pattern of genetic influence shared across multiple traits.

### Bootstrap aggregation and co-appearance estimation

To assess robustness, we performed bootstrap aggregation with *B* = 500 resamples. In each bootstrap iteration, signal and noise SNPs were resampled separately from their original sets, with equal sample sizes (80% subsample), and all subsequent steps (SVD completion, covariance estimation, factor extraction, and rotation) were recomputed from scratch.

Each iteration produced a list of gene–trait clusters *L*_*b*_. We repeat from the resampling B times and obtain a collection of cluster list {*ℒ*_b_}. Based on the multiple gene-trait clusters {*ℒ*_b_}, we propose to aggregate the cluster lists using a co-appearance graph. Consider a graph whose nodes denote SNPs and traits. For two nodes v_i_ and v_j_, we define the weight for the edge connecting v_i_ and v_j_

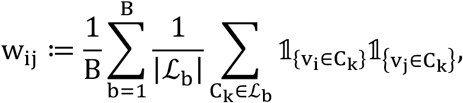

Where C_k_ denotes the gene-trait clusters in the list *ℒ*_b_ obtained from the b-th bootstrap sample.

Let C ∈ ℝ^T×T^ be the normalized trait–trait co-appearance matrix, where C_ij_ measures how often traits i and j co-appear across repeated PathGPS runs. Traits are assigned to K clusters G_1_, …, G_k_.

### Clustering and dimensional reduction

Trait geometry was visualized using Uniform Manifold Approximation and Projection (UMAP) applied to the normalized trait–trait co-appearance matrix with correlation distance.

Discrete genetic modules were identified using spectral co-clustering (implemented via sklearn.cluster.SpectralCoclustering) applied to the bipartite SNP×trait co-appearance matrix. We evaluated cluster resolutions (*K* = 4, 5, 6). When the effective number of non-empty clusters was four (i.e., clusters containing both non-missing SNPs and traits), stability metrics indicated the most reproducible solution.

Clustering stability was assessed using adjusted Rand indices and consensus matrices across repeated runs with different random seeds.

### Stability and permutation testing

Clustering stability was evaluated by repeating spectral co-clustering across 20 independent runs with different random seeds. Reproducibility was quantified using adjusted Rand indices and consensus matrices.

We conducted permutation tests to assess statistical significance of the recovered clusters. Let C ∈ ℝ^T×T^ be the normalized trait–trait co-appearance matrix, where C_ij_ measures how often traits i and j co-appear across repeated PathGPS runs. Traits are assigned to K clusters G_1_, …, G_k_.

The global test statistic was defined as

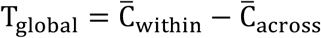

Where 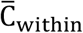 and 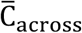 denote average of C_ij_ where traits i and j are in the same cluster and are in different clusters.

The individual test statistic was defined as

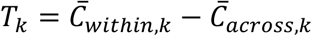

Where 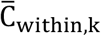 and 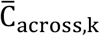 denote average of *C*_*i*j_ where traits *i* and *j* are in the cluster *k* and traits *i* are in the cluster *k* while traits *j* are in different clusters.

We permute trait labels while preserving cluster sizes (|*G*_1_|, …, |*G*_*k*_|), recompute 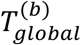 and 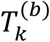 for *b* = 1, …, *B*, and compute cluster discrepancy score as

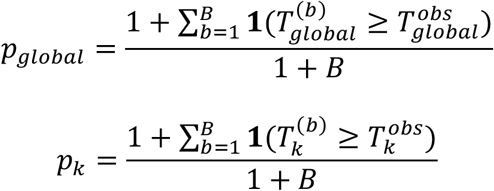

Trait labels were permuted while preserving cluster sizes, and the cluster discrepancy scores were computed from 1,000 permutations. Lower scores indicate greater differences between clusterings, and are therefore preferred.

### Simulation

We simulate SNP--trait summary associations under two generative schemes that control (i) sample overlap across traits and (ii) environmental correlation among traits.

For each dataset we draw *n* = 1500 individuals and *p* = 5000 SNPs. Genotypes are simulated with per--SNP minor allele frequencies *MAF* ∼ *Unif* (0.1, 0.5) and then centered and standardized.

We create sparse loading matrices *U* ∈ ℝ^*p*×*r*^ (*SNP* → *genetic mediators*) and *V* ∈ ℝ^*M*×*r*^ (*mediators* → *traits*) with columnwise sparsity (nonzero entries ∼ *N*(0, 1)). Unless stated otherwise we use *M* = 30 traits and *r* = 13 latent genetic factors.

We generate *s*_*env*_ latent environmental factors 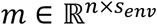 with covariance Σ_*e*_. Two structures are used:

i. compound symmetry:

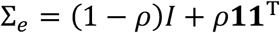

controls the strength *ρ* of shared environment.
ii. low rank:

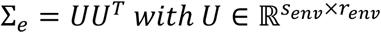

Trait--specific environmental loadings are 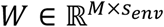 (sparse).

In our simulator, all traits share the same genotype matrix *X* (sample overlap). Latent mediators are *M*_*lat*_ = *XU* + *ϵ*_*M*_,and traits are *Y* = *M*_*lat*_*V*^*T*^ + *mW*^*T*^ + *ϵ*_F_,with *ϵ*_*M*_, *ϵ*_F_ i.i.d.Gaussian noise. Marginal SNP--trait effects are 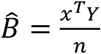

We study four scenarios, each replicated 50 times:

- B1 (no env. corr.): simulator with shared *X, s*_*env*_ = 3, compound symmetry with *ρ* = 0 (environment present but uncorrelated across latent envs).
- B2 (Overlap, moderate env. corr.): shared *X*, compound symmetry with *ρ* = 0.3.
- B3 (Overlap, strong env. corr.): shared *X*, compound symmetry with *ρ* = 0.8.
- LowRank (Overlap, low-rank env.): shared *X*, low-rank Σ_*e*_ with *r*_*env*_ = 2.

The main text reports the setting with *M* = 30 traits and *r* = 8 genetic factors (the real number of genetic factors generated in the simulation is 13, but set the model to discover 8 factors); additional (*M, r*) combinations appear in the Appendix.

We compare PathGPS with SVD, hard-thresholded SVD (SVD_HT), sparse principal components (SPC), maximum likelihood factor analysis (MLFA), and genetic factor analysis (GFA). Below we specify, for each baseline, the input matrix, preprocessing, and factor dimension settings.

### SVD and SPC baselines (matrix-factorization on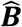)

SVD and SPC operate directly on the marginal association matrix 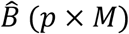. For SVD, we compute the truncated singular value decomposition of 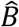 and retain the top *r* right singular vectors *M* × *r* as the estimated trait loadings. For SVD_HT, we apply elementwise hard-thresholding to 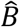 (setting entries with 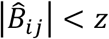 to 0, with *z* = 4 in our implementation) and then run truncated SVD, retaining the top r components. For SPC, we start from the top *r* SVD loading vectors and enforce sparsity via thresholding (approximating an *L*1 ‘sumabs’ constraint); each loading is re-normalized to unit *L*2 norm after thresholding. All three methods use the same component count r as PathGPS in the corresponding simulation setting.

MLFA_4 / MLFA_8 (factor analysis on an LDSC-like trait correlation matrix). MLFA is applied to a trait-trait matrix analogous to an LDSC genetic correlation matrix. In simulation, we construct this matrix as the correlation across SNPs of the columns of 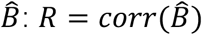), where corr(.) denotes Pearson correlation computed across SNPs. We then fit maximum-likelihood factor analysis to R using scikit-learn’s FactorAnalysis. We evaluate two factor numbers in the main-text setting: MLFA_8 (k = 8) and MLFA_4 (k = 4), representing a correctly specified and a deliberately under-specified model size. After fitting, we apply varimax rotation to the loading matrix to improve interpretability.

### GFA (Genetic Factor Analysis on 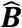 with standard errors)

GFA is fit using the R package GFA (Jean Morrison’s implementation), called from Python via rpy2. GFA takes as input 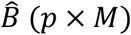 and an accompanying standard-error matrix *S* (*p* × *M*). In the simulator, we estimate the standard error for each trait *j* as 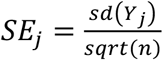 and tile these SE values across SNPs to form S. Unless otherwise stated, we use the package default priors, initialization, and convergence criteria. When a maximum number of factors is required, we set kmax to match the target factor dimension for comparability (e.g., kmax = 8 in the main-text setting).

We evaluate recovery using three metrics: (i) Discovered factors: number of true latent factors correctly identified; (ii) Extraneous factors: number of spurious or noise-driven factors; and (iii) Mismatch score: alignment error between estimated and true loadings, computed after matching estimated factors to true factors using an optimal assignment based on factor-wise correlations.

### Real-data analysis: GenomicSEM genome-wide factor modeling

To benchmark PathGPS against established multivariate genetic modeling, we conducted a real-data analysis using the same GWAS data we use for PathGPS analysis. We assembled trait-specific summary statistics files (one per phenotype) and ensured a consistent trait naming and ordering across all downstream steps to prevent inadvertent misalignment between LD Score Regression (LDSC) outputs and structural equation model (SEM) fitting. All analyses were performed in R using the GenomicSEM package.

### Summary-statistics harmonization and LDSC genetic covariance estimation

We first harmonized and quality-controlled each GWAS summary statistics file using GenomicSEM’s munge() procedure, restricting SNPs to the HapMap3 reference panel. We only use the necessary (must-input) information from the GWAS data set, so we did not apply filters on imputation quality and minor allele frequency (we do not include the MAF and INFO in our dataset). For each trait, sample size was taken from the per-SNP N column when available. For binary traits, we specified trait-specific sample prevalence and population prevalence parameters to place SNP effects on the liability scale, whereas continuous traits were treated with missing prevalence values. We then estimated the multivariate SNP-heritability and genetic covariance matrix across traits using multivariable LDSC (ldsc()), based on European LD reference files.

### Trait inclusion criteria and model stability

Because low-powered traits can destabilize the genetic covariance structure and induce non-positive definite (non-PSD) covariance estimates, we implemented a priori QC filter based on SNP-heritability z-scores. Specifically, we computed the heritability z-statistic as 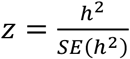 and retained only traits with *z* ≥ 2. This filtering step removed traits whose genetic signal was insufficient for reliable multivariate modeling. We additionally excluded one dementia/ADR phenotype (ADRD) prior to LDSC due to downstream instability in the multivariate covariance estimation. LDSC was re-run on the retained trait set to obtain the final genetic covariance matrix used for SEM estimation.

### S-matrix smoothing prior to exploratory factor inspection

In practice, multivariate LDSC genetic covariance matrices can be slightly non-PSD due to sampling variability and estimation noise. To facilitate exploratory factor inspection and stabilize subsequent model building, we smoothed the LDSC genetic covariance matrix using a nearest positive-definite projection (nearPD in the Matrix package), applied to the LDSC S-matrix (genetic covariance) while preserving the original covariance scale (i.e., without forcing a correlation matrix). The smoothed covariance matrix was used only as a diagnostic aid to support factor-structure exploration; confirmatory estimation used the LDSC covariance structure as input to GenomicSEM.

### Factor-number selection and confirmatory genome-wide model specification

We selected the number of latent genetic factors empirically by fitting and inspecting a sequence of candidate structures with 2, 3, 4, and 5 factors. For each candidate, we assessed overall model fit (including comparative fit indices reported by GenomicSEM) and the interpretability and stability of factor loadings. This iterative procedure was used to refine trait-to-factor assignments and avoid degenerate factors (e.g., factors defined by weak or unstable loadings) while maintaining parsimony. Based on these comparisons, we specified a five-factor confirmatory model that separated major domains of shared genetic liability and treated educational attainment as an independent latent dimension to prevent it from distorting psychopathology factor structure.

### Genome-wide confirmatory factor model fitting (DWLS)

The final five-factor genome-wide model was estimated using GenomicSEM’s usermodel() with diagonally weighted least squares (DWLS) and unit-variance identification for latent variables (std.lv = TRUE). In this model, factors were allowed to correlate freely to capture broad genetic sharing across domains. The fitted model produced standardized factor loadings and residual variances for each trait, enabling decomposition of each trait’s SNP-heritability into variance explained by the latent genetic factors versus trait-specific background components. We saved the fitted model object for downstream comparison and visualization, and used the final factor model as a reference structure for evaluating PathGPS-derived trait clustering and genetic relationship recovery.

### Final five-factor model used in real data

The final confirmatory model comprised five correlated latent genetic factors: an internalizing/affective factor capturing shared liability among post-traumatic stress and neuroticism-related traits (with an additional neurological trait retained based on QC), a psychotic/compulsive spectrum factor capturing shared liability among schizophrenia, bipolar disorder, and obsessive–compulsive disorder, a neurodevelopmental factor capturing shared liability among ADHD, ASD, and Alzheimer’s disease within the retained trait set, a mood/compulsivity factor capturing shared liability between major depression and obsessive–compulsive symptoms, and an educational attainment factor modeled as a separate latent dimension. This specification reflects the empirically observed loading patterns across candidate models while prioritizing model stability under multivariate LDSC uncertainty.

### Real-Data Comparison with GFA

To benchmark PathGPS against an established latent factor approach, we applied Genetic Factor Analysis (GFA) to the same set of real GWAS summary statistics and compared the inferred trait–factor structure. For each trait, we used genome-wide SNP-level summary statistics containing chromosome, base-pair position, rsID, effect and non-effect alleles, marginal effect estimates (*β*), standard errors, sample size, and p-values.

Prior to model fitting, summary statistics were subjected to standard quality control. SNPs with non-positive or missing standard errors were removed. To ensure a unique representation of each variant within a trait, duplicated rsIDs were resolved by retaining the entry with the smallest p-value. All traits were then harmonized to a common effect-allele orientation using a reference GWAS (major depressive disorder), and strand-ambiguous variants (A/T and C/G) were excluded. Effect sizes were flipped as needed to maintain consistent allele alignment across traits.

### Construction of the SNP × trait matrix

After harmonization, we constructed a wide *SNP* × *traits* matrix by merging summary statistics across traits using a union strategy. To balance SNP coverage against data completeness, we retained only variants observed in at least eight of the fifteen traits. For each retained SNP–trait pair, we computed z-scores as the ratio of the marginal effect estimate to its standard error. This resulted in a dense z-score matrix spanning tens of thousands of variants and all analyzed traits.

Trait-specific effective sample sizes were summarized using the median per-SNP sample size for each trait, providing a stable estimate consistent with recommended GFA practice for heterogeneous GWAS designs.

### Genetic Factor Analysis

GFA was fit to the *SNP* × *trait* z-score matrix using the standard GFA likelihood framework, which models genetic associations as arising from a low-rank latent factor structure with trait-specific loadings. In the primary real-data analysis, we supplied z-scores and per-trait effective sample sizes, and allowed the model to infer latent factors under the assumption of independent residual noise. Estimation of nuisance correlation due to sample overlap was explored but not required for the primary trait–factor comparison, as our focus was on the recovered factor loadings rather than trait–trait covariance estimates.

The output of interest was the estimated trait–factor loading matrix 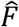, which summarizes how each trait loads onto the inferred genetic factors. This matrix provides a direct basis for comparison with the pathway-based trait structure recovered by PathGPS.

### Role of GFA in the comparison

GFA serves as a representative latent factor model that captures shared genetic architecture through global, low-rank structure. In contrast, PathGPS identifies pathway-level co-appearance patterns that need not conform to a single low-rank factorization. By applying both methods to the same harmonized real GWAS data, we assess how pathway-based representations differ from and complement factor-analytic summaries of cross-trait genetic sharing.

### Negative control analysis: hair colour (UK Biobank)

To evaluate whether PathGPS can recover genetically meaningful multi-trait structure without being driven by arbitrary or unrelated phenotypes, we performed a negative-control analysis by adding a trait expected to be largely independent of psychiatric, cognitive, and neurodegenerative genetic architecture. Specifically, we included Hair colour (natural, before greying): Blonde (UK Biobank phenotype code 1747_1) as an additional trait and repeated the full PathGPS pipeline, including all preprocessing, signal–noise decomposition, covariance adjustment, bootstrapping, and co-clustering, without modifying any hyperparameters.

UK Biobank GWAS summary statistics for the hair-colour phenotype were obtained using the UKBB GWAS Imputed v3 documentation and metadata (UKBB GWAS Imputed v3 Google Sheet). Because UK Biobank summary statistics are indexed by an internal variant identifier, we used the UKBB “List of variants used in GWAS, with annotations” file (variants.tsv.bgz) as a reference map to recover genomic coordinates and rs identifiers. We merged the hair-colour GWAS results with this variant reference to obtain harmonized SNP fields (CHR, BP, and rsID/SNP) and then appended hair colour as an additional trait to the multi-trait SNP–trait matrix. This negative-control trait was loaded immediately after the original GWAS datasets and then processed identically to all other traits, ensuring that any observed effects reflect the PathGPS model behavior rather than differential preprocessing.

We used this negative-control setting to assess robustness in two complementary ways. First, we examined whether the inclusion of hair colour alters trait-module structure among the original phenotypes (i.e., whether major clusters and their stability are preserved). Second, we checked whether hair colour is assigned as a coherent module on its own or spuriously embedded into psychiatric/neurodegenerative clusters. Together, these diagnostics provide an internal specificity check, testing whether PathGPS separates unrelated genetic variation from the shared structure of interest.

### Statistical environment

All analyses were conducted in Python using standard numerical and machine-learning libraries, with PLINK used for LD calculations and scikit-learn used for spectral co-clustering. We provide full computational details—including preprocessing scripts, parameter settings, and reproducible workflows—through the publicly accessible PathGPS GitHub repository (https://github.com/Hxw-w/PathGPS_NeuroDisorders).

## Data Availability

All genome-wide association study (GWAS) summary statistics used in this study are publicly available from the original study consortia and data repositories.

GWAS summary statistics for attention deficit hyperactivity disorder (ADHD), autism spectrum disorder (ASD), bipolar disorder (BIP), major depressive disorder (MDD), obsessive–compulsive disorder (OCD), obsessive–compulsive symptoms (OCS), schizophrenia (SCZ), and post-traumatic stress disorder (PTSD) were obtained from the Psychiatric Genomics Consortium and are available at https://pgc.unc.edu/for-researchers/download-results/.

GWAS summary statistics for neuroticism were obtained from the Complex Trait Genetics group and are available at https://cncr.nl/ctg/.

GWAS summary statistics for Alzheimer’s disease and related dementias (ADRD) were obtained from the National Institute on Aging Genetics of Alzheimer’s Disease Data Storage Site (NIAGADS), dataset NG00075, available at https://dss.niagads.org/datasets/ng00075/.

GWAS summary statistics for amyotrophic lateral sclerosis (ALS) were obtained from the NHGRI–EBI GWAS Catalog (https://www.ebi.ac.uk/gwas/), accession ID GCST90027164, corresponding to European ancestry meta-analyses.

GWAS summary statistics for educational attainment (EA) were obtained from the Social Science Genetic Association Consortium (SSGAC) at http://www.thessgac.org/data. Access to these data is subject to the consortium’s terms of use, which encourage responsible use of the data. The analyses used association results for all SNPs passing quality-control filters from autosomal, X-chromosome, and dominance GWAS meta-analyses that exclude research participants from 23andMe.

All analyses in this study were restricted to GWAS summary statistics derived from individuals of European ancestry, as reported in the original studies.

## Code Availability

The full analytical pipeline used in this study, including data preprocessing, LD-based imputation, modeling, clustering, and figure generation, is available in the PathGPS Psychology project repository at: https://github.com/Hxw-w/PathGPS_NeuroDisorders. This repository contains the original end-to-end workflow, intermediate outputs, and scripts used to generate the results reported in this manuscript.

A modular implementation of the PathGPS framework, developed subsequently for general research use, is available at: https://github.com/Hxw-w/pathgps. The pathgps package provides reusable components for preprocessing, modeling, and downstream analysis, along with demonstration scripts and a Jupyter notebook illustrating typical outputs.

All analyses were conducted using Python 3.10 and PLINK v1.9.0-b.7.11 (64-bit, 19 Aug 2025). Specific package dependencies are listed in the respective repositories.

