## Supplementary Notes for "Background covariance adjustment distills shared genetic architecture across neurodevelopmental and neurodegenerative disorders"

Top genes preserved across original and negative-control clustering. Traits shown in each cluster represent the intersection of traits that co-clustered both in the original PathGPS analysis and after adding the negative-control trait (hair color), ensuring that listed modules are highly stable.

| Cluster | Traits | Top genes |
|---|---|---|
| **1** | ADHD, EA, BIP | KIZ/KIZ-AS1, supported by cross-disorder GWAS (ADHD–ASD–OCD; ADHD–ASD); L351–L359; SOX11, neuronal differentiation; linked to schizophrenia 39†L1469–L1472; TRAF3, associated with ADHD, bipolar, and depression. |
| **2** | panic, ocd, asd | PVALB, parvalbumin interneuron deficits in ASD; optogenetic modulation relieves OCD-like behavior. |
| **3** | ptsd, scz, neuro, ocs | PDE4B, implicated in schizophrenia and PTSD (GWAS and functional mouse models 62†L11–L39); PATJ, rare synaptic variants in schizophrenia and ASD 68†L74–L82; NFASC, altered expression and myelination abnormalities in schizophrenia. |
| **4** | adrd, als | CSMD2, complement-regulator gene implicated in AD endophenotypes 51†L31–L39; HIVEP3, AD-associated variant (rs10493098) and increased expression in patient blood. |

**Supplementary Table 1.** Most genes without psychiatric links were filtered out during literature review. Key genes such as PDE4B, NFASC, and PVALB appear repeatedly in large-scale GWAS or functional models, supporting their plausibility. In the Alzheimer's cluster, both CSMD2 and HIVEP3 have emerging evidence from AD-related imaging genetics or APOE-independent risk. This supports PathGPS's ability to yield replicable, biologically meaningful genes for each latent trait module.

**Stability of SNP–trait co-clustering**

To assess the robustness of PathGPS biclusters, we performed 20 runs of biclustering (K=5) and computed consensus co-assignment matrices. As shown in Figure bellow, both SNP and trait clustering exhibit high consistency. The majority of SNPs have pairwise co-assignment >0.6 across replicates, with many reaching >0.9. Trait consensus is even sharper, with nearly block-diagonal structure indicating near-perfect assignment stability.

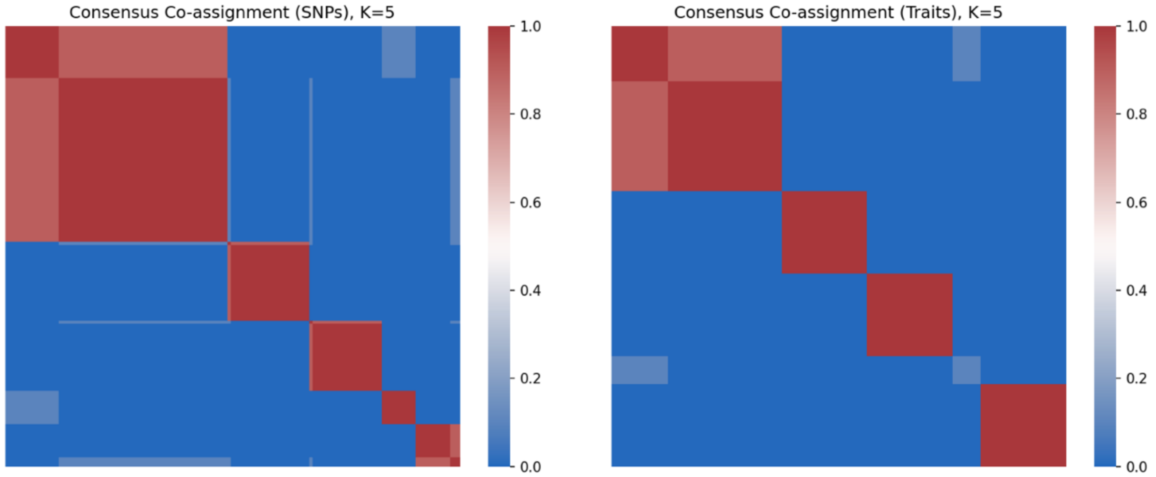


**Supplementary Figure 1**. Consensus co-assignment matrices for SNPs (left) and traits (right) across 20 bootstrap runs at K= 5. Warmer colors indicate greater consistency of assignment.

**Negative control: clustering remains stable with non-psychiatric trait**

To test specificity, we added a negative control—GWAS summary statistics for hair color (blonde)—to the trait set and reran PathGPS. As summarized in Table 5, the original trait clusters remain largely intact. Hair color does not form connections with any psychiatric modules, and all core trait groupings (e.g., PTSD–SCZ–neuroticism; ADHD–EA–BIP) are preserved.

| Cluster | Original traits | With hair color added |
|---|---|---|
| 1 | alz, neuro, ptsd, scz, ocs | alz, neuro, ptsd, scz, ocs |
| 2 | adhd, adrd, als, ea, mdd, bip | adhd, adrd, als, ea, mdd, bip, hair color |
| 3 | an | an |
| 4 | panic, asd, ocd | panic, asd, ocd |

**Spplementary Table 2**: Comparison of trait cluster membership before and after adding hair color as a negative control.

**Additional Simulation Results**

Additional simulations were perfromed for examining the performance of PathGPS. Panels evaluating PathGPS against baseline methods across multiple data-generating conditions. The main text reports the (30 traits, 8 latent factors, 13 real latent factors) condition, as it represents a balanced, high-signal scenario. For completeness, we include here the additional settings: (30 traits, 6 factors, 10 real latent factors), (50 traits, 10 factors, 15 real latent factors). Across all scenarios, PathGPS consistently identifies the correct latent factors with fewer extraneous factors and lower mismatch error compared to SVD, hard-thresholded SVD, SPC, and MLFA variants.

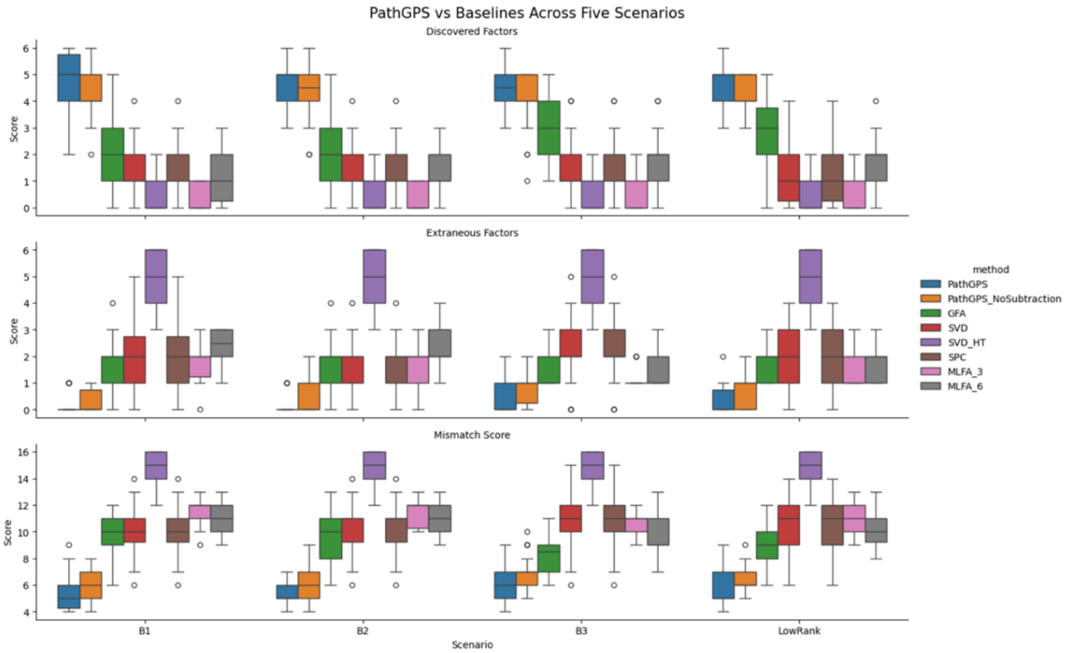


**Supplementary Figure 2**. Simulation results for the (30 traits, 6 factors) setting. Performance is evaluated across five scenarios (A, B1, B2, B3, LowRank) using three metrics: discovered factors, extraneous factors, and mismatch score.

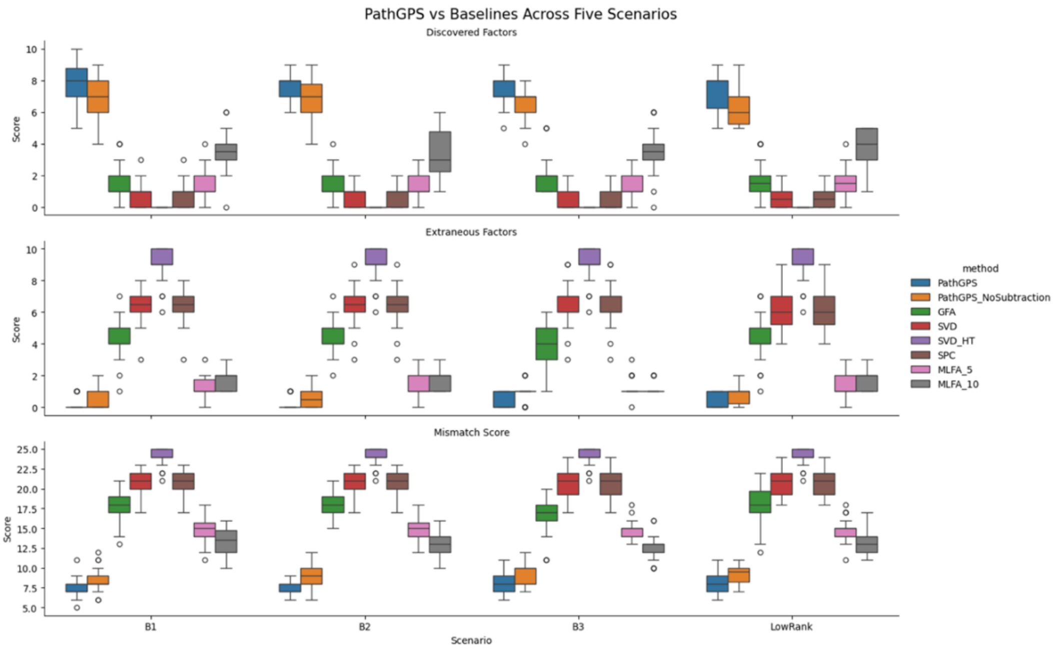


**Supplemenary Figure 3**. Simulation results for the (50 traits, 10 factors, 15 real factors) setting. Even in higher dimensional regimes, PathGPS avoids over-factoring and maintains stable mismatch performance.

### Top genes consistently assigned to clusters

To identify biologically meaningful genes, we focused on SNPs that remained in the same clusters with and without the negative control, and annotated them via Ensembl VEP. The resulting gene lists, assumed to reflect stable cluster-level associations, are shown in Supplemnetary Table 1. For each, we searched literature to validate trait relevance. Each gene is listed with references (where available) connecting it to the psychiatric or neurological traits defining its cluster. Empty entries indicate no relevant peer-reviewed evidence was found linking that gene to the cluster’s traits.

Cluster 1 (ADHD, Educational Attainment, Bipolar)

*KIZ-AS1*: Cross-disorder GWAS meta-analyses implicate the KIZ/KIZ-AS1 locus in ADHD and autism (https://pubmed.ncbi.nlm.nih.gov).
*SOX11*: A neurodevelopmental transcription factor, has been linked to psychiatric disorders (https://pmc.ncbi.nlm.nih.gov; https://mdpi.com).
*KIZ*: Same evidence as for KIZ-AS1 (https://pubmed.ncbi.nlm. nih.gov).
*TRAF3*: The TRAF3 locus shows pleiotropic association across ADHD,bipolar disorder and depression (https://medrxiv.org).
*UTP4:* No peer-reviewed studies linking UTP4 (CIRH1A) to ADHD, education or bipolar (https://ncbi.nlm.nih.gov).
*FTLP18*: No literature associating FTLP18 with ADHD, education or bipolar traits.
*RPS15AP1:* No evidence linking RPS15AP1 to ADHD, educational attainment or bipolar disorder.
TCF12-DT: No publications connecting TCF12-DT to ADHD, education or bipolar disorder.

Cluster 2 (Panic Disorder, OCD, Autism)

*KHDRBS3*: No direct evidence linking KHDRBS3 to panic disorder, OCD, or ASD.
*SF3A3*: No studies connect the splicing-factor gene SF3A3 to panic disorder, OCD, or autism.
*PPP1R13B*: No reported associations between PPP1R13B (ASPP1) and panic, OCD, or autism.
*PVALB*: No evidence linking LINC01748 to panic disorder, OCD, or ASD. PVALB encodes parvalbumin, a calcium-binding protein of fast-spiking GABA interneurons. PV interneuron deficits are implicated

in autism (https://frontiersin.org); PV-knockout mice show autism-like behaviors, and optogenetic activation of PV neurons alleviates grooming in OCD models (https://nature.com).
*VPS4A*: No specific links found between VPS4A and panic disorder,OCD, or autism.

Cluster 3 (PTSD, Schizophrenia, Neuroticism, OC Symptoms)
*PDE4B*: PDE4B lies in a schizophrenia locus (1p31) and interacts with DISC1 (https://sciencedirect.com); multiple studies link PDE4B variants to schizophrenia and PTSD-like behaviors (https://jneurosci.org).
*NFASC:* NFASC (Neurofascin) regulates myelinated axons; post-mortem analyses show NFASC down-regulation in schizophrenia (https://pmc.ncbi.nlm.nih.gov;https://frontiersin.org).

Cluster 4 (Alzheimer's Disease & Related Dementias, ALS)
*CSMD2*: CSMD2 implicated in AD via imaging-genetics GWAS (https://neurosci.cn); encodes complement-pathway regulator expressed in brain (https://genHIVEP3  org).
*HIVEP3*: Variant rs10493098 C in HIVEP3 associated with AD risk,especially APOE-ε4 negative (https://alz-journals.onlinelibrary.wiley.com); expression elevated in AD patients.

### Supplementary Table 3. GWAS datasets analyzed in this study

| Category | Trait (abbreviation) | Full name | Primary GWAS reference |
|---|---|---|---|
| Psychiatric disorders | ADHD | Attention deficit hyperactivity disorder | Demontis et al., Nature Genetics (2023); Psychiatric Genomics Consortium |
| | ASD | Autism spectrum disorder | Grove et al., Nature Genetics (2019); Psychiatric Genomics Consortium |
| | SCZ | Schizophrenia | Trubetskoy et al., Nature (2022); Psychiatric Genomics Consortium |
| | BIP | Bipolar disorder | O'Connell et al., Nature (2024); Psychiatric Genomics Consortium |
| | PTSD | Post-traumatic stress disorder | Nievergelt et al., Nature Genetics (2024); Psychiatric Genomics Consortium |
| | MDD | Major depressive disorder | PGC MDD Working Group, Cell (2024) |
| | PANIC | Panic disorder | Forstner et al., Molecular Psychiatry (2019); Psychiatric Genomics Consortium |

| | | | |
|---|---|---|---|
| | OCD | Obsessive–compulsive disorder | Strom et al., Nature Genetics (2025); Psychiatric Genomics Consortium |
| | OCS | Obsessive–compulsive symptoms | Strom et al., Molecular Psychiatry (2024); Psychiatric Genomics Consortium |
| | AN | Anorexia nervosa | Watson et al., Nature Genetics (2019); PGC Eating Disorders Working Group |
| Personality / cognitive traits | NEURO | Neuroticism | Nagel et al., Nature Genetics (2018) |
| | EDU | Educational attainment | Okbay et al., Nature Genetics (2022); SSGAC |
| Neurodegenerative outcomes | ALZ | Alzheimer's disease | Bellenguez et al., Nature Genetics (2019) |
| | ADRD | Alzheimer's disease and related dementias | Shade et al., Nature Genetics (2024) ; NIAGADS Consortium; large-scale meta-analyses curated in NIAGADS (dataset NG00075) |
| | ALS | Amyotrophic lateral sclerosis | van Rheenen et al., Nature Genetics (2021); NHGRI–EBI GWAS Catalog (ID GCST90027164) |
